# Sex-Difference of Associations between Cigarette Smoking and Myocardial Fibrosis: The Multi-Ethnic Study of Atherosclerosis

**DOI:** 10.1101/2025.07.29.25332410

**Authors:** Elie W. Akl, Ralph Zeitoun, Omar Chehab, Haiou Li, Vinithra Varadarajan, Colin O. Wu, Alain G. Bertoni, Karol E. Watson, David A. Bluemke, Bharath Ambale-Venkatesh, João A.C. Lima

**Author notes:** Equal contribution (Elie W. Akl, Ralph Zeitoun). Address of correspondence: Elie Akl, MD, Division of Cardiology, Brigham and Women’s Hospital, 70 Francis St, Boston, MA 02115.

## Abstract

**Background:** Extracellular volume (ECV) and native T1 time (nT1) are markers of interstitial-myocardial-fibrosis (IMF) by cardiac MRI (CMR) and are associated with CV events, heart failure and death. However, the association between smoking and IMF at the population level has not been explored.

**Objectives:** This study investigated the relationship between smoking and IMF by CMR, as well as the sex differences of this association in the MESA cohort.

**Methods:** A total of 2118 participants (53% women) had data on smoking between 2000 and 2012 and ECV(%) and nT1 (ms) at exam-5 (2010-2012). We constructed participant-specific trajectories of average cigarettes per day (ACPD) based on linear interpolation to estimate ACPD between exams 1 through 5 (ACPD_1-5_). We explored the associations of smoking status at Exam-5, ACPD at Exam-5, ACPD_1-5_ and temporal change in smoking status, with ECV and nT1 using multivariable analysis.

**Results:** Current smoking status was associated with increased markers of IMF in women but not in men, ECV% (□ =2% p=<0.001 vs. 0.5%; p=0.2) and nT1 (□ =28ms; p=<0.001 vs. 3ms; p=0.5) in women vs. men, respectively. A higher ACPD_1-5_ was associated with increased ECV% (□ =0.1%; p<0.001) and nT1 (□ =1ms; p=0.003) in women but not in men. Similarly, higher ACPD at Exam-5 was associated with higher ECV% in women vs. men. Sex interaction ascertained prior to stratification was statistically significant.

**Conclusion:** In this population study, cigarette smoking was associated with a higher prevalence of IMF in females but not in males, indicating differences in the pathophysiology of CVD by sex.

## INTRODUCTION

Myocardial fibrosis is characterized by the accumulation of extracellular matrix in the myocardium and has been associated with several cardiovascular diseases including myocardial infarction, as well as dilated and hypertrophic cardiomyopathies. When it comes to the identification of myocardial fibrosis, there are various imaging modalities available (such as echocardiography, single-photon emission computed tomography, PET, and CT), however contrast-enhanced magnetic resonance imaging is the most accurate method for the detection of myocardial fibrosis. Late-gadolinium enhancement (LGE) and T1 mapping are commonly used techniques for measuring myocardial scarring and the extracellular volume fraction of the myocardium, respectively. T1 mapping has been used to identify and quantify diffuse and infiltrative interstitial fibrosis. Contrast-enhanced T1 mapping allows for the calculation of extracellular volume (ECV) expansion which is a marker of myocardial tissue remodeling and is associated with adverse collagen deposition ^1–3^. In addition, prolonged native T1 measurements are also useful as indices of diffuse myocardial fibrosis in patients with structurally altered extracellular matrix architecture, in association with cardiomyopathy ^2–4^.

Smoking is one of the main modifiable risk factors for cardiovascular disease and a main accelerator of cardiovascular ageing. Studies looking directly at the link between cigarette smoking and myocardial remodeling have consolidated the unfavorable effects of smoking on the structure and function of the cardiovascular system ^5^. However, there are few studies that evaluated the impact of smoking on interstitial myocardial fibrosis. In this regard, a greater pack-year smoking history has been associated with higher rates of atrial fibrosis in patients undergoing coronary artery bypass ^6^. In addition, in a cohort of patients with dilated cardiomyopathy undergoing CMR, smoking was associated with a higher incidence of myocardial scar and fatal ventricular arrhythmias ^7^.

To our knowledge, no prior studies have evaluated the effect of gender and smoking on interstitial fibrosis in a community population. Several animal studies have shown that cigarette smoking is associated with myocardial fibrosis and myocardial dysfunction through increased oxidative stress ^8–10^.

In this study, we used CMR to evaluate the impact of cigarette smoking on the presence of interstitial myocardial fibrosis in a population without baseline CVD. In addition, we sought to evaluate sex differences in the association between cigarette smoking and myocardial fibrosis.

## METHODS

### Study Population

The Multi-Ethnic Study of Atherosclerosis (MESA) is a prospective, observational cohort study that included 6814 men and women with no prior history of cardiovascular disease from four racial/ethnic groups (White, Black, Hispanic/Latino, Chinese) and recruited from six states over two years between 2000-2002. A detailed study design has been described previously ^11^. The Institutional Review Boards approved the study protocol of participating institutions, and each participant signed informed consent before recruitment^12^. Baseline examination (Exam 1) was conducted between 2000 and 2002 and then participants were followed up and data were collected over four subsequent follow-up visits (Exam 2 [2002-2004]; Exam 3 [2004-2005]; Exam 4 [2004-2005]; and Exam 5 [2010-2012]). MESA participants who have smoking data available at Exam 1 and Exam 5 and have received a cardiac MRI at Exam 5 with available native T1 time (ms) or ECV (%) were included. Those with myocardial infarction or congestive heart failure by Exam 5 were excluded (**Supplemental Figure 1**).

### Cigarette Smoking

Cigarette smoking data in MESA was collected through a questionnaire administered at each exam. Smoking status was assessed through the following questions: 1) “Have you smoked at least 100 cigarettes in your lifetime?”, 2) “Have you smoked during the last 30 days?”. If the participant answered No to the first question, then they are classified as “Never Smoker”. If they answered Yes to the first question, then based on the second question they are classified as either “Current Smoker” if they answered Yes or “Former Smoker” if they answered No. Smoking intensity was assessed based on the following question “On average, about how many cigarettes a day do/did you smoke?”.

For each participant with at least 3 available data measurements regarding the average number of cigarettes smoked per day at Exam 1, Exam 5, and any of the remaining exams (2-4), the cumulative smoking exposure between Exam 1 and Exam 5 was calculated. To explore the cumulative exposures and time-trends of smoking, we considered the age-based time-trend analysis described in Wu and Tian ^13^. Since the data on the number of cigarettes per day is right-skewed and zero-inflated, we calculated subject-specific trajectories of smoking exposure using the linear interpolation introduced in Wu and Tian ^13^. The area under the subject-specific trajectories for the i-th participant’s cigarettes per day for the past 10 years is calculated by 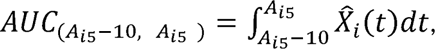 where *A*_*i*5_ is the age of i-th participant at exam 5. The data were analyzed using R version 4.2.3.

To measure temporal changes in smoking exposure, we categorized participants based on their smoking status at Exam 1 and Exam 5 into four categories: never smokers at exams 1 and 5 are labeled as “Never Smoker”; former smokers at exams 1 and 5 are labeled “Former Smoker for More Than 10 years”; former smokers at exam 5 and never or current smokers at exam 1 are labeled “Former Smoker for Less Than 10 years”; and current smokers at exams 1 and 5 are labeled as “Smoker for More than 10 years”.

### CMR Protocol

The Cardiac MRI protocol was described previously^14^. In brief, CMR was performed using 1.5 T scanners (Avanto and Espree, Siemens Medical Systems; Signa LX, GE Healthcare) with a 6-channel anterior phased-array torso coil and posterior coil elements. Interstitial myocardial fibrosis was evaluated using the single breath-hold modified Look-Locker (MOLLI) inversion recovery sequence. Pre- and post-contrast injection mid-LV short-axis T1 maps were obtained. QMass (Research version, Leiden, Netherlands) was used to analyze the images. A region of interest (ROI) was manually delineated around the core myocardium on each T1 map (pre- and post-contrast) to determine the myocardial T1 time. If a myocardial scar was present, it was included in the ROI. Greater diffuse myocardial fibrosis has been linked to lower post-contrast T1 times. The partition coefficient was derived by plotting the myocardium’s 1/T1 timings against the blood pool and measuring the slope of the resulting linear regression line. ECV = Partition Coefficient*(1-hematocrit/100) was used to compute the extracellular volume fraction (ECV). Considering the limited availability of hematocrit lab values, we used a synthetic ECV based on the blood’s longitudinal relaxation rate. This has been shown to have a high correlation to conventional ECV and CVD outcomes ^15^

### Statistical Analysis

Clinical characteristics and demographics from the evaluation during exam 5 were used for analysis. Categorical variables are summarized as counts and relative frequencies, while continuous variables were summarized as mean ± SD. Independent sample t-test was used to compare column means and chi-square test was used to compare proportions. Linear regression was used to evaluate the association between native T1 and ECV and each of smoking status at exam 5, reported average number of cigarettes smoked per day at exam 5, 10-year cumulative smoking exposure between exam 1 and exam 5 and smoking habit changes between exam 1 and exam 5. Model 1 covariates include age, sex, and race. Model 2 includes Model 1 covariates in addition to highest level of education, BMI, systolic blood pressure, use of anti-hypertensive medications, LDL level, use of lipid-lowering medications, diabetes status and estimated GFR. Model 3 includes Model 2 covariates in addition to Mean Agatston Coronary Artery Calcium Score and the presence of late gadolinium enhancement (LGE) by cardiac MRI. These confounders are known to influence the development of myocardial fibrosis and affect parametric mapping as previously described in multiple studies^16,17^. Statistical significance was defined as p<.05. Interaction with sex was tested for all analyses, and subsequent stratification by sex was performed when interaction was found to be significant. Unstandardized beta coefficients are presented. Statistical analyses were performed using SPSS Statistics version 28.

## RESULTS

### Population characteristics

A total of 2118 participants (53% women, mean age 68.6 years) had cigarette smoking data and had undergone CMR analysis for myocardial fibrosis. Of those, 1309 participants had available ECV data, and 2118 participants had available native T1 data. Participants with a history of MI or CHF by the time of CMR acquisition (at Exam 5) were excluded from the analysis. The baseline characteristics of the study population stratified by sex are provided in **Table 1**.

**Table 1:**
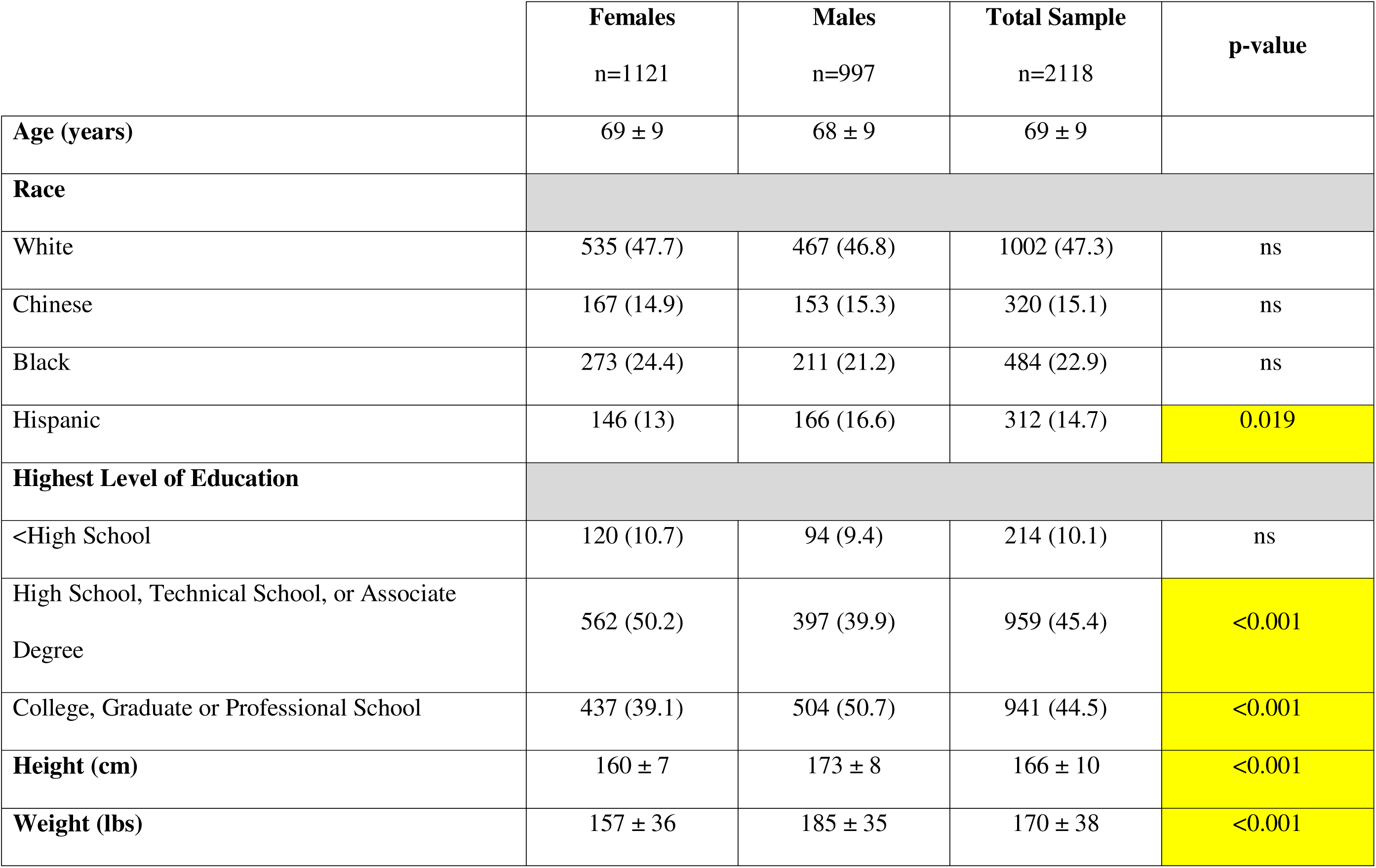
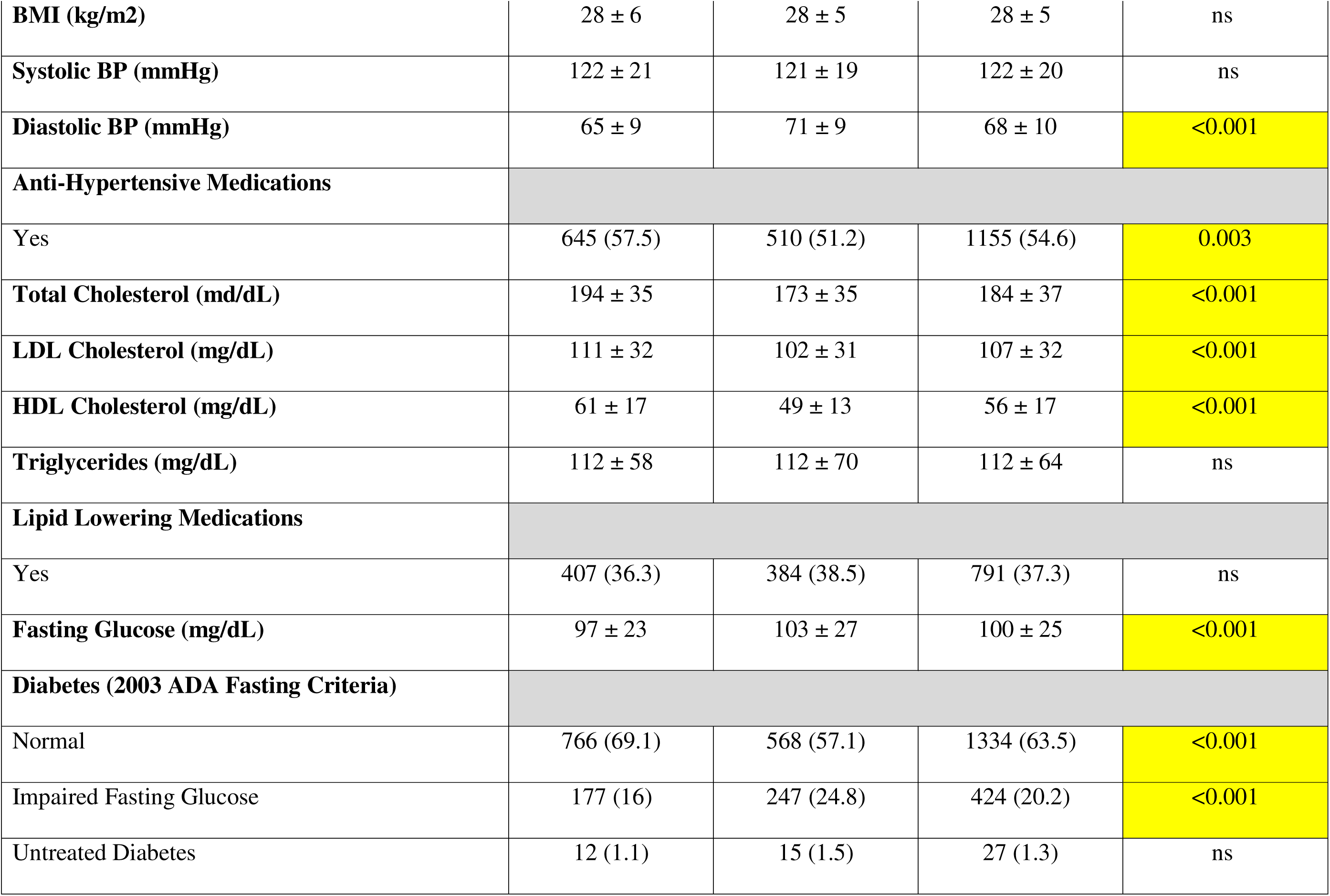
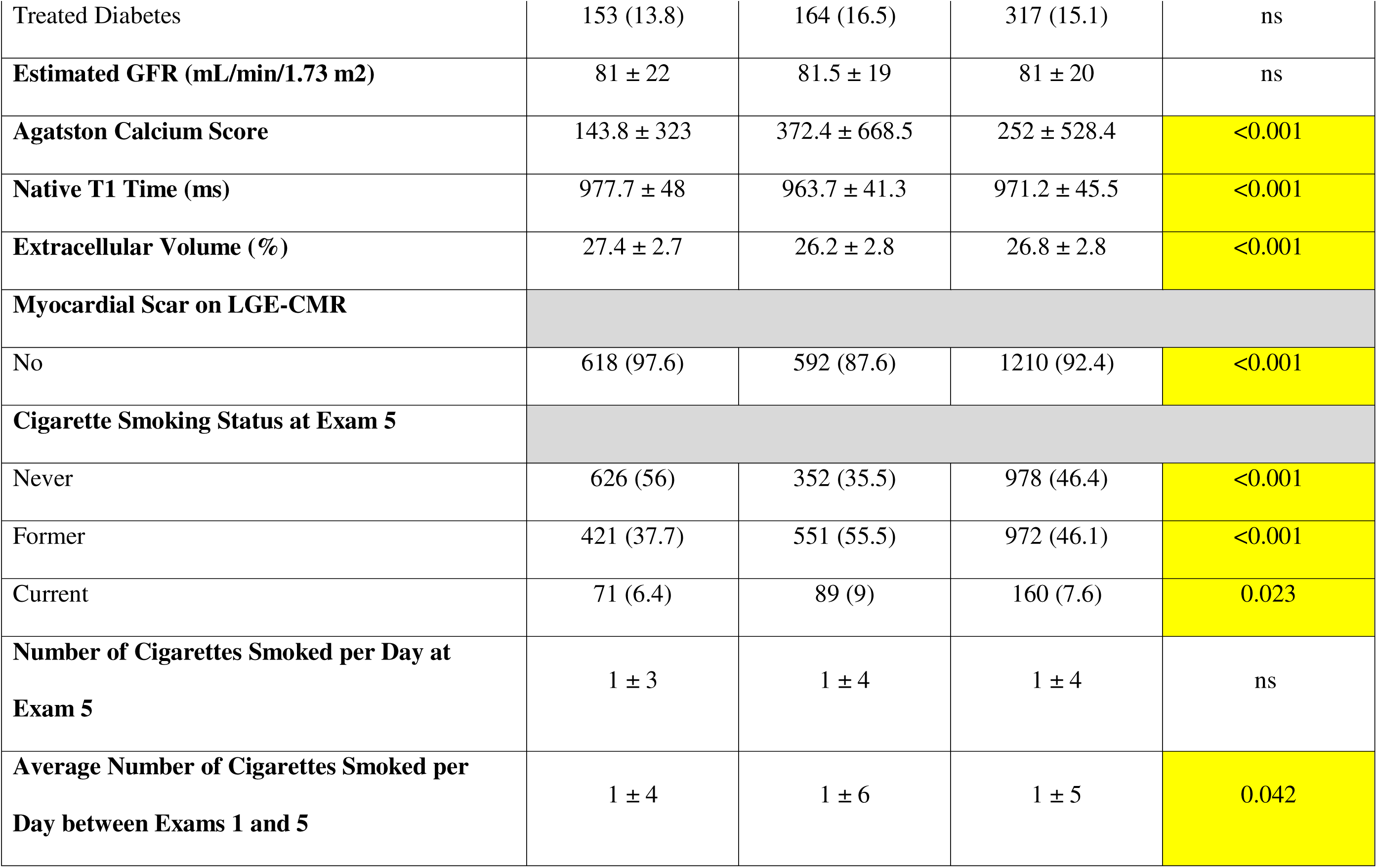
Demographics of Participants at Exam 5.

### Smoking status and myocardial fibrosis

**Tables 2 and 3** show that current smoking status at Exam 5 was significantly associated with higher ECV and native T1 time, respectively, independent of demographic characteristics and traditional cardiovascular risk factors. There was a significant interaction between sex and current smoking status for ECV and with both former and current smoking status for native T1 time. After stratifying by sex, current smoking status was independently associated with 2% increase in ECV% and a ∼28ms increase in native T1 time in females but not in males in the unadjusted and fully adjusted analysis (p<0.001 for both). There was no significant association between former smoking status and ECV or native T1 time in male or female participants.

**Table 2:** Multivariable linear regression models for association and sex interaction of cigarette smoking status with ECV (%) at Exam 5.

| Total Sample |  | Unadjusted |  | Model 1 |  | Model 2 |  | Model 3 |  |
| --- | --- | --- | --- | --- | --- | --- | --- | --- | --- |
| | | $\beta$ (95% CI) | p | $\beta$ (95% CI) | p | $\beta$ (95% CI) | p | $\beta$ (95% CI) | p |
| Smoking Status at Exam 5 | Never Smoker | ref | ref | ref | ref | ref | ref | ref | ref |
|  | Former Smoker | 0.2 (-0.2-0.5) | 0.3 | 0.3 (-0.03-0.7) | 0.077 | 0.3 (-0.1-0.6) | 0.098 | 0.3(-0.03-0.7) | 0.08 |
|  | Current Smoker* | 0.8(0.1-1.5) | **0.01** | 1.3 (0.6-1.9) | **<.001** | 1.2 (0.6-1.8) | **<.001** | 1.1 (0.5-1.7) | **<.001** |

\* Significant interaction with sex; $p = 0.029$

| Male |  | Unadjusted |  | Model 1 |  | Model 2 |  | Model 3 |  |
| --- | --- | --- | --- | --- | --- | --- | --- | --- | --- |
| | | $\beta$ (95% CI) | p | $\beta$ (95% CI) | p | $\beta$ (95% CI) | p | $\beta$ (95% CI) | p |
| Smoking<br>Status at<br>Exam 5 | Never Smoker | ref | ref | ref | ref | ref | ref | ref | ref |
|  | Former Smoker | 0.5 (-0.03-1) | 0.06 | 0.3 (-0.2-0.8) | 0.279 | 0.3 (-0.2-0.8) | 0.266 | 0.3 (-0.2-0.8) | 0.251 |
|  | Current Smoker | 0.5(-0.4-1.3) | 0.3 | 0.7 (-0.2-1.5) | 0.112 | 0.5 (-0.3-1.4) | 0.21 | 0.5 (-0.4-1.3) | 0.287 |

| Female |  | Unadjusted |  | Model 1 |  | Model 2 |  | Model 3 |  |
| --- | --- | --- | --- | --- | --- | --- | --- | --- | --- |
| | | $\beta$ (95% CI) | p | $\beta$ (95% CI) | p | $\beta$ (95% CI) | p | $\beta$ (95% CI) | p |
| Smoking<br>Status at<br>Exam 5 | Never Smoker | ref | ref | ref | ref | ref | ref | ref | ref |
|  | Former Smoker | 0.3 (-0.2-0.8) | 0.3 | 0.1 (-0.4-0.7) | 0.573 | 0.1 (-0.4-0.7) | 0.595 | 0.2 (-0.3-0.7) | 0.514 |
|  | Current Smoker | 1.9 (0.9-2.8) | **<.001** | 2 (1.1-3) | **<.001** | 2 (1.1-3) | **<.001** | 2 (1-3) | **<.001** |

**Table 3:** Multivariable linear regression models for association and sex interaction of cigarette smoking status with Native T1 time (ms) at Exam 5.

| Total Sample |  | Unadjusted |  | Model 1 |  | Model 2 |  | Model 3 |  |
| --- | --- | --- | --- | --- | --- | --- | --- | --- | --- |
| | | $\beta$ (95% CI) | p | $\beta$ (95% CI) | p | $\beta$ (95% CI) | p | $\beta$ (95% CI) | p |
| Smoking Status at Exam 5 | Never Smoker | ref | ref | ref | ref | ref | ref | ref | ref |
|  | Former Smoker* | -4 (-9-2) | 0.2 | -0.24 (-61-59) | 0.938 | -0.1 (-6-5) | 0.972 | 0.2 (-5-6) | 0.954 |
| | Current Smoker± | 9 (-2-19) | 0.09 | 13 (34-232) | **0.011** | 13 (3-23) | **0.009** | 13 (3-23) | **0.011** |

\*Significant interaction with sex; \* $p=0.009$ ; $\pm p=0.006$

| Male |  | Unadjusted |  | Model 1 |  | Model 2 |  | Model 3 |  |
| --- | --- | --- | --- | --- | --- | --- | --- | --- | --- |
| | | $\beta$ (95% CI) | p | $\beta$ (95% CI) | p | $\beta$ (95% CI) | p | $\beta$ (95% CI) | p |
| Smoking<br>Status at<br>Exam 5 | Never Smoker | ref | ref | ref | ref | ref | ref | ref | ref |
|  | Former Smoker | -0.6 (07-6) | 0.9 | -3 (-10-5) | 0.48 | -2 (-9-5) | 0.65 | -2 (-9-6) | 0.66 |
|  | Current Smoker | 2 (-9-15) | 0.7 | 2 (-10-15) | 0.7 | 2 (-10-14) | 0.73 | 2 (-10-14) | 0.765 |

| Female |  | Unadjusted |  | Model 1 |  | Model 2 |  | Model 3 |  |
| --- | --- | --- | --- | --- | --- | --- | --- | --- | --- |
| | | $\beta$ (95% CI) | p | $\beta$ (95% CI) | p | $\beta$ (95% CI) | p | $\beta$ (95% CI) | p |
| Exam 5 | Never Smoker | ref | ref | ref | ref | ref | ref | ref | ref |
|  | Former Smoker | -0.1 (-9-8) | 0.9 | -1(-1-8) | 0.822 | -2 (-10-7) | 0.6831 | -1 (-10-8) | 0.829 |
|  | Current Smoker | 25 (8-41) | **0.003** | 26 (10-43) | **0.002** | 30 (13-46) | **<.001** | 30 (13-46) | **<.001** |

### Dose-dependent effect of smoking and myocardial fibrosis

**Tables 4 and 5** show that a higher reported average number of cigarettes per day at Exam 5 was significantly associated with higher ECV (β=1; p<0.001) and native T1 time (β=1; p=0.011), respectively, independent of demographic characteristics and traditional cardiovascular risk factors. Higher calculated mean number of cigarettes per day was also significantly associated with higher ECV (β=0.1; p<0.001) and native T1 time (β=0.5; p<0.001). There was a significant interaction between sex and both the reported average number of cigarettes per day at Exam 5 and the calculated mean number of cigarettes per day for ECV, however, the interaction with sex was only significant with the calculated mean number of cigarettes per day for native T1 time. When stratified by sex, both the reported average number of cigarettes per day at Exam 5 (β=0.1; p<0.001) and the calculated mean number of cigarettes per day (β=0.1; p<0.001) were significantly associated with higher ECV, and the calculated mean number of cigarettes per day was associated with higher native T1 (β=1; p=0.003) in females but not in males in the fully adjusted analysis (Figure 1).

**Figure 1.**
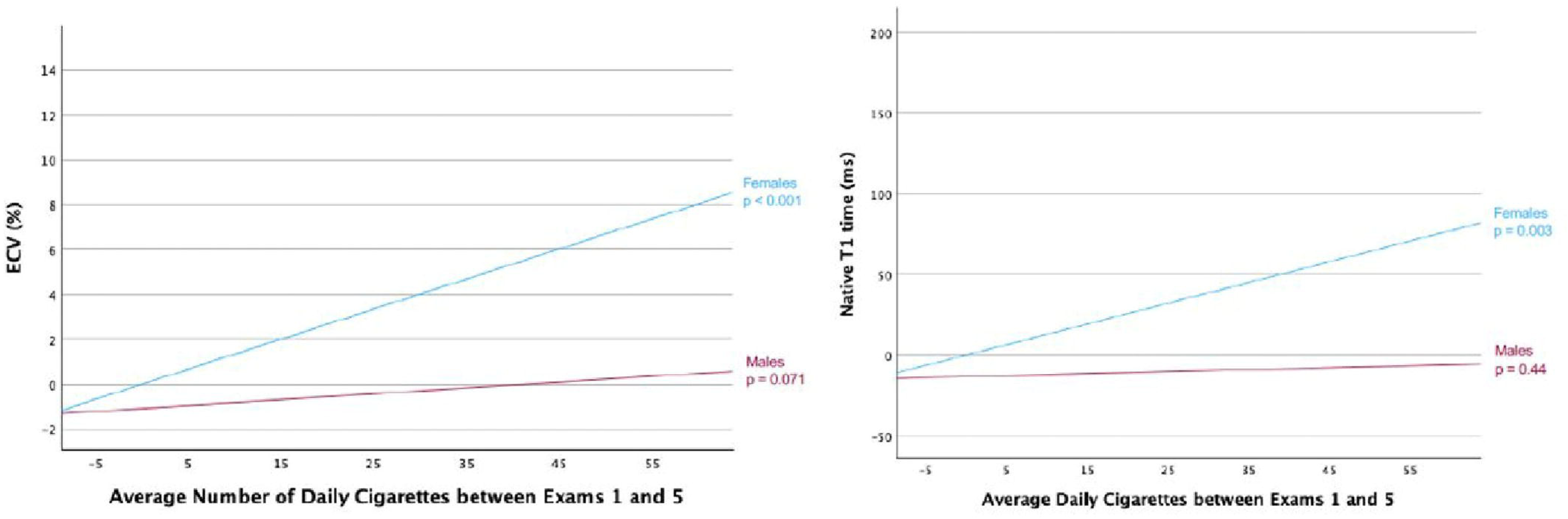
Sex Differences in Smoking-Related Myocardial Fibrosis. Partial regression plots demonstrating sex-stratified association between average number of daily cigarettes and markers of interstitial myocardial fibrosis by cardiac MRI after controlling for demographic characteristics and traditional cardiac risk factor

**Table 4:** Multivariable linear regression models for association and sex interaction of average number of cigarettes smoked per day (at Exam 5, and calculated average between Exams 1 and 5) and ECV (%)

| Total Sample | Model 1 |  | Model 2 |  | Model 3 |  |
| --- | --- | --- | --- | --- | --- | --- |
| | $\beta$ | p | $\beta$ | p | $\beta$ | p |
| Average Number of Cigarettes smoked per day at Exam 5* | 0.1 (0.04-0.12) | **<.001** | 0.1 (0.04-0.12) | **<.001** | 0.1 (0.03-0.12) | **<.001** |
| Calculated Average Number of Cigarettes smoked per day between Exam 1 & 5<sup>±</sup> | 0.1 (0.02-0.1) | **<0.001** | 0.1 (0.02-0.1) | **<0.001** | 0.1 (0.02-0.1) | **<0.001** |

\* ± Significant interaction with sex; \* $p=0.016$ ; $\pm p<0.001$

| Male | Model 1 |  | Model 2 |  | Model 3 |  |
| --- | --- | --- | --- | --- | --- | --- |
| | $\beta$ | p | $\beta$ | p | $\beta$ | p |
| Average Number of Cigarettes smoked per Day at Exam 5 | 0.1 (0-0.1) | 0.058 | 0.04 (-0.01-0.1) | 0.091 | 0.04 (-0.01-0.1) | 0.156 |
| Calculated Average Number of Cigarettes smoked per day between Exam 1 & 5 | 0.02 (-0.002-0.06) | 0.069 | 0.03 (-0.002-0.06) | 0.071 | 0.03 (-0.004-0.06) | 0.091 |

| Female | Model 1 |  | Model 2 |  | Model 3 |  |
| --- | --- | --- | --- | --- | --- | --- |
| | $\beta$ | p | $\beta$ | p | B | p |
| Average Number of Cigarettes smoked per day at Exam 5 | 0.1 (0.1-0.2) | **<.001** | 0.1 (0.1-0.2) | **<.001** | 0.1 (0.1-0.2) | **<.001** |
| Calculated Average Number of Cigarettes smoked per day between Exam 1 & 5 | 0.1 (0.1-0.2) | **<0.001** | 0.1 (0.1-0.2) | **<0.001** | 0.1 (0.1-0.2) | **<0.001** |

**Table 5:**
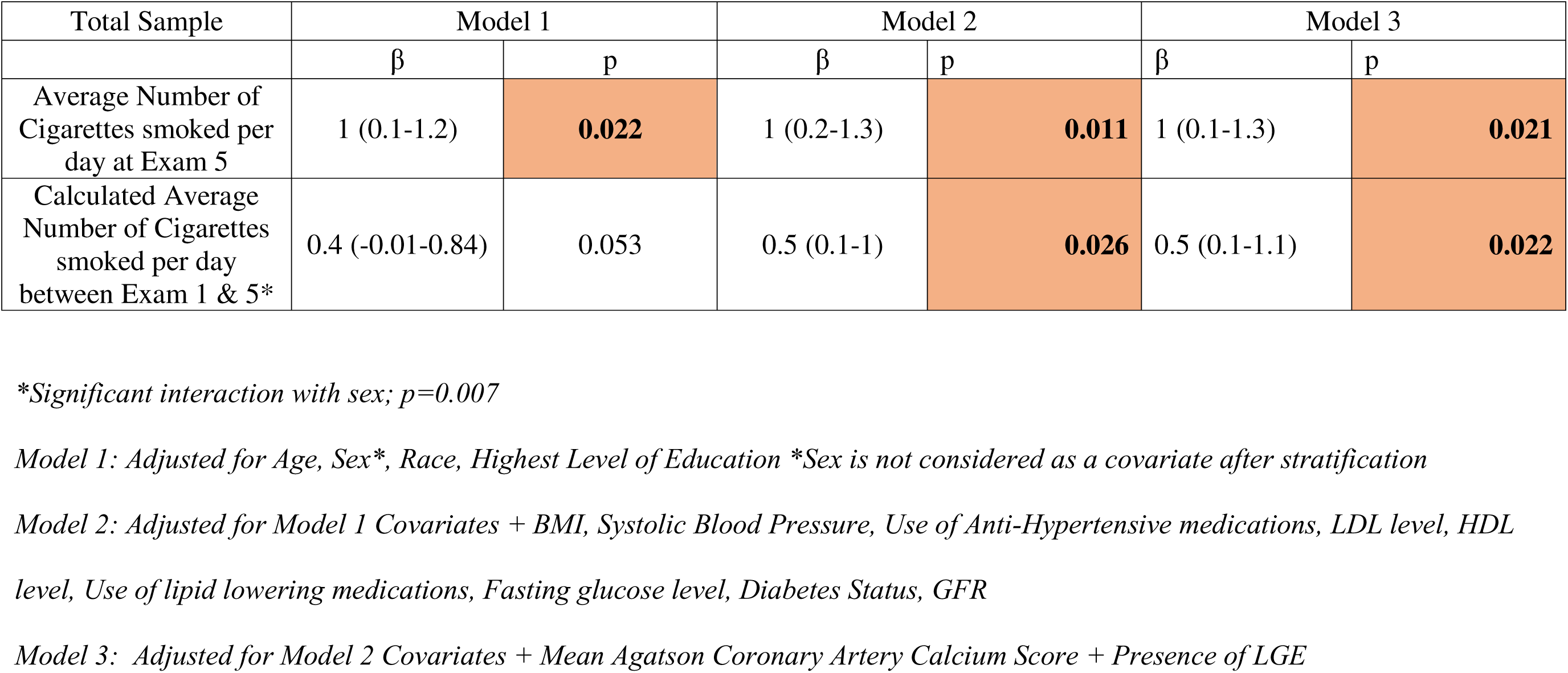
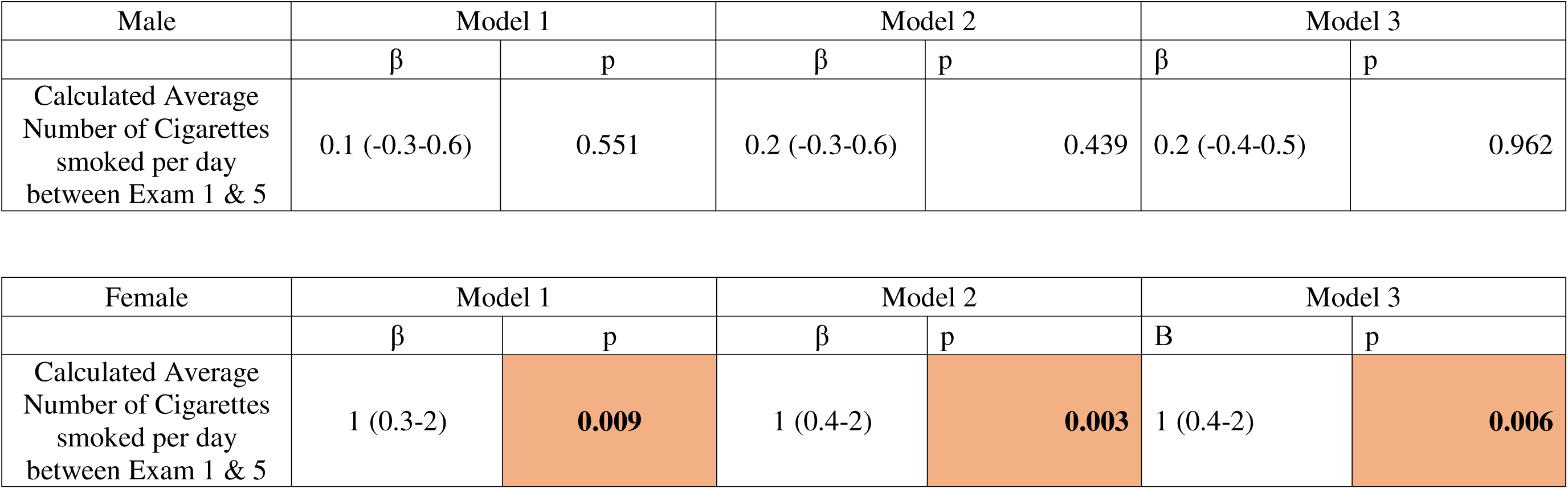
Multivariable linear regression models for association and sex interaction of average number of cigarettes smoked per day (at Exam 5, and calculated average between Exams 1 and 5) and Native T1 time (ms)

### Temporal change in smoking habit and myocardial fibrosis

As shown in **Tables 6 and 7**, being a smoker for more than 10 years was associated with a 2 % increase in ECV % ( p<0.001) and 29 ms increase in native T1 time ( p<0.001), in females but not in males. However, smoking cessation for more than 10 years was not associated with increase in ECV or native T1 time. Moreover, smoking cessation for less than 10 years was significantly associated with a 0.7% increase in ECV (p=0.012) but no change in native T1 time and the former association is lost when stratified by sex.

**Table 6:** Multivariable linear regression models for association and sex interaction of change in cigarette smoking status between Exams 1 and 5 and ECV (%)

| Total Sample | Model 1 |  | Model 2 |  | Model 3 |  |
| --- | --- | --- | --- | --- | --- | --- |
| | $\beta$ | p | $\beta$ | p | $\beta$ | p |
| Never Smoker | ref | ref | ref | ref | ref | ref |
| Former Smoker for More Than 10 Years | 0.2 (-0.2-0.6) | 0.285 | 0.2 (-0.2-0.5) | 0.355 | 0.2 (-0.2-0.6) | 0.273 |
| Former Smoker For Less Than 10 Years | 0.7 (0.2-1.3) | 0.012 | 0.7 (0.2-1.3) | 0.011 | 0.7 (0.1-1.3) | 0.014 |
| Smoker for More Than 10 Years* | 1.6 (0.9-2.2) | <0.001 | 1.4 (0.8-2.1) | <0.001 | 1.4 (0.7-2) | <0.001 |

*\*Significant interaction with sex; $p=0.031$*

| Males | Model 1 |  | Model 2 |  | Model 3 |  |
| --- | --- | --- | --- | --- | --- | --- |
| | $\beta$ | p | $\beta$ | p | $\beta$ | p |
| Never Smoker | ref | ref | ref | ref | ref | ref |
| Former Smoker For More Than 10 Years | 0.1 (-0.4-0.7) | 0.595 | 0.2 (-0.4-0.7) | 0.537 | 0.2 (-0.3-0.7) | 0.481 |
| Former Smoker For Less Than 10 Years | 0.8 (0.03-1.6) | 0.042 | 0.8 (-0.01-1.5) | 0.054 | 0.7 (-0.1-1.5) | 0.071 |
| Smoker for More Than 10 Years | 0.9 (-0.03-1.8) | 0.059 | 0.8 (-0.1 -1.7) | 0.096 | 0.7 (-0.3-1.6) | 0.156 |

| Females | Model 1 |  | Model 2 |  | Model 3 |  |
| --- | --- | --- | --- | --- | --- | --- |
| | $\beta$ | p | $\beta$ | p | $\beta$ | p |
| Never Smoker | ref | ref | ref | ref | ref | ref |
| Former Smoker For More Than 10 Years | 0.1 (-0.5-0.6) | 0.851 | 0.03 (-0.5-0.6) | 0.916 | 0.01 (-0.5-0.6) | 0.798 |
| Former Smoker For Less Than 10 Years | 0.5 (-0.4-1.4) | 0.285 | 0.5 (-0.4-1.4) | 0.24 | 0.5 (-0.4-1.4) | 0.25 |
| Smoker for More Than 10 Years | 2.3 (1.3-3.3) | <0.001 | 2.3 (1.3-3.3) | <0.001 | 2.3 (1.2-3.3) | <0.001 |

**Table 7:**
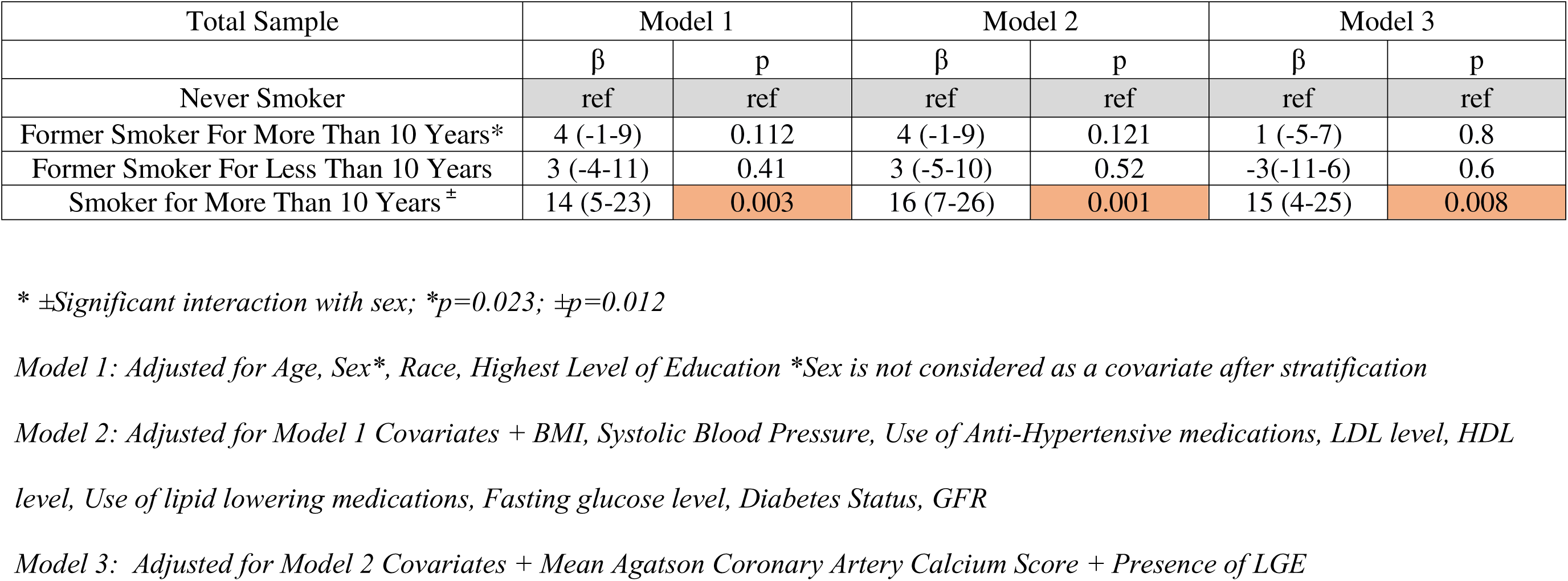
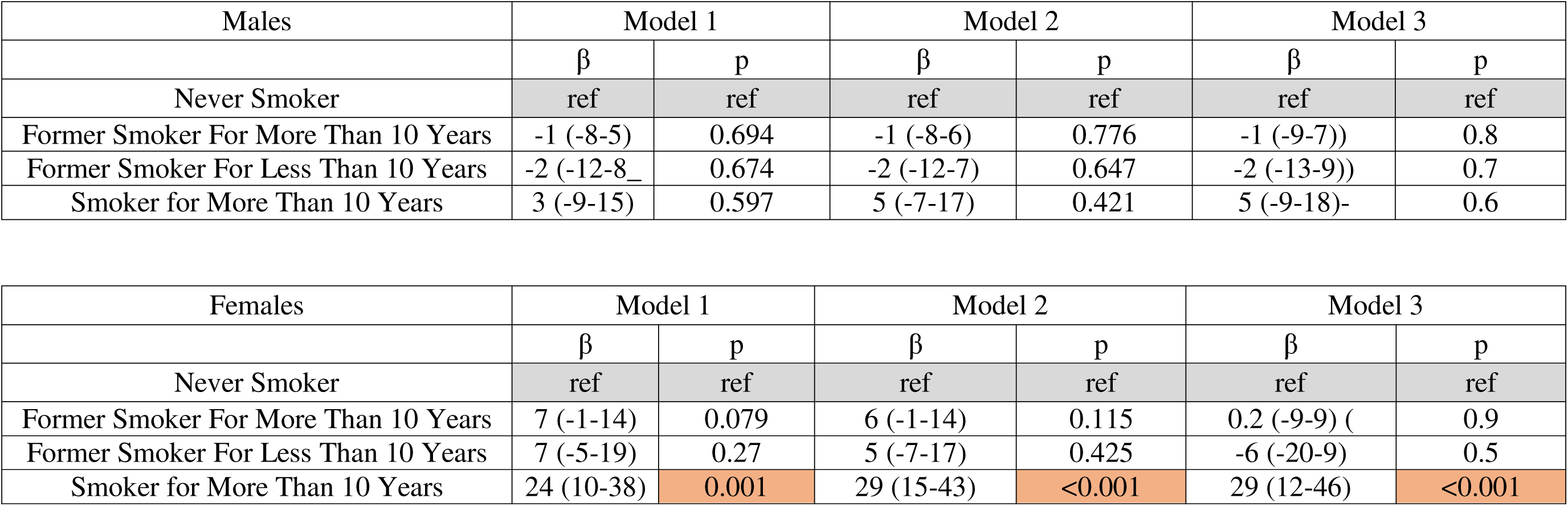
Multivariable linear regression models for association and sex interaction of change in cigarette smoking status between Exams 1 and 5 and Native T1 (ms)

| Total Sample | Model 1 |  | Model 2 |  | Model 3 |  |
| --- | --- | --- | --- | --- | --- | --- |
| | $\beta$ | p | $\beta$ | p | $\beta$ | p |
| Never Smoker | ref | ref | ref | ref | ref | ref |
| Former Smoker For More Than 10 Years* | 4 (-1-9) | 0.112 | 4 (-1-9) | 0.121 | 1 (-5-7) | 0.8 |
| Former Smoker For Less Than 10 Years | 3 (-4-11) | 0.41 | 3 (-5-10) | 0.52 | -3(-11-6) | 0.6 |
| Smoker for More Than 10 Years <sup>±</sup> | 14 (5-23) | 0.003 | 16 (7-26) | 0.001 | 15 (4-25) | 0.008 |

\* ± Significant interaction with sex; \* $p=0.023$ ; $\pm p=0.012$

| Males | Model 1 |  | Model 2 |  | Model 3 |  |
| --- | --- | --- | --- | --- | --- | --- |
| | $\beta$ | p | $\beta$ | p | $\beta$ | p |
| Never Smoker | ref | ref | ref | ref | ref | ref |
| Former Smoker For More Than 10 Years | -1 (-8-5) | 0.694 | -1 (-8-6) | 0.776 | -1 (-9-7)) | 0.8 |
| Former Smoker For Less Than 10 Years | -2 (-12-8) | 0.674 | -2 (-12-7) | 0.647 | -2 (-13-9)) | 0.7 |
| Smoker for More Than 10 Years | 3 (-9-15) | 0.597 | 5 (-7-17) | 0.421 | 5 (-9-18)- | 0.6 |

| Females | Model 1 |  | Model 2 |  | Model 3 |  |
| --- | --- | --- | --- | --- | --- | --- |
| | $\beta$ | p | $\beta$ | p | $\beta$ | p |
| Never Smoker | ref | ref | ref | ref | ref | ref |
| Former Smoker For More Than 10 Years | 7 (-1-14) | 0.079 | 6 (-1-14) | 0.115 | 0.2 (-9-9) ( | 0.9 |
| Former Smoker For Less Than 10 Years | 7 (-5-19) | 0.27 | 5 (-7-17) | 0.425 | -6 (-20-9) | 0.5 |
| Smoker for More Than 10 Years | 24 (10-38) | 0.001 | 29 (15-43) | <0.001 | 29 (12-46) | <0.001 |

## DISCUSSION

In this study, we found that current smoking status in females but not in males is associated with higher markers of myocardial fibrosis by CMR in a population with no prior CVD events. In addition, our analysis revealed a significant dose-dependent effect between smoking burden at the time of the cardiac MRI and cumulative smoking exposure over a 10-year period prior to CMR acquisition and the prevalence of myocardial fibrosis in female participants. Importantly also, we found that the association between smoking and myocardial fibrosis was lost for participants who stopped smoking, particularly if cessation occurred more than 10 years before fibrosis assessment. These findings align with emerging evidence of sex-specific cardiovascular responses to smoking and underscores the utility of advanced imaging in detecting subclinical myocardial changesFew studies have investigated the relationship between smoking and interstitial myocardial fibrosis. In a cross-sectional study of 381 patients without known cardiovascular disease, current smoking status was associated with elevated ECV when compared to former and non-smokers (0.25%, 95%CI 0.24–0.26% vs. 0.24%, 95%CI 0.24–0.26%, p<0.01) ^18^. Another study of 239 patients with type 2 diabetes mellitus showed that active smoking was associated with a 2% increase in ECV when compared with non-smokers ^19^. Our results corroborate these findings, as we demonstrated that current smoking status was associated with greater ECV and longer native T1 time. However, when stratified by sex we found that this association was mainly driven by a ∼2% ECV increase among female participants who were current smokers when compared to nonsmoker MESA participants.

Several studies have established a significant association between smoking intensity and increased risk of CVD mortality^20–23^. Using the MESA cohort, Nance et. al showed that the average intensity of smoking at baseline is associated with worse CVD outcomes in current smokers^24^. Furthermore, research has shown that even relatively low levels of fine particulate exposure from cigarette smoke are sufficient to induce adverse biological responses and increase the risk of cardiovascular disease mortality. The exposure-response relationship between cardiovascular disease mortality and fine particulate matter is relatively steep at low levels of exposure and flattens out at higher exposures ^25^. Our results showed similar dose-dependent relationship between smoking exposure and myocardial fibrosis by CMR. Increasing smoking intensity reflected by the reported number of cigarettes smoked per day was associated with higher markers of fibrosis. Moreover, higher cumulative exposure of smoking over a 10 year period prior to CMR acquisition was associated with higher fibrosis markers.

Smoking cessation is widely recommended to reduce CVD burden. However, the timeline for the reduction of cardiovascular risk after quitting smoking in comparison to never smokers remain undefined. The ASCVD Risk Estimator Plus states that the CVD risk of former smokers becomes similar to those who never smoked after 5 years^26^. Duncan et al. showed that among heavy smokers (≥20 pack-years), those who successfully quit smoking exhibited a significantly reduced risk of CVD within the first 5 years following cessation, in comparison to their counterparts who continued smoking (hazard ratio of 0.61). In addition, it took approximately 10 to 15 years following smoking cessation for former heavy smokers to no longer exhibit a statistically significant increase in CVD risk when compared to individuals who had never smoked^27^. Moreover, in a recent study, Ding et al. showed that smoking cessation was significantly associated with lower risk of heart failure, however the residual risk persisted for a few decades after smoking cessation^28^. Our findings support these results, as we showed that the association between smoking and interstitial fibrosis by CMR was no longer significant in participants who stopped smoking for more than 10 years prior to fibrosis assessment.

Several studies have consistently demonstrated that at baseline, females exhibit higher native T1 values and ECV than males, suggesting inherent physiological differences in myocardial structure and function between the sexes. One study found that females have higher myocardial perfusion, blood volume, and ECV compared to males, both at rest and during stress conditions, indicating that sex is an independent contributor to these parameters beyond other physiological factors^29^. Another study highlighted that female sex is associated with higher native T1 values, even after adjusting for variables such as age, heart rate, and left ventricular mass index, further supporting the notion of intrinsic sex-based disparities in myocardial tissue composition^30^.

Previous studies have assessed the sex-differentiated effect of smoking on the development of cardiovascular disease. The Copenhagen Heart study found that smoking was associated with a higher risk for all ACSs in women when compared to men ^31^. Similarly, it has been shown that female smokers are more likely to develop STEMI when compared to male smokers aged <65 years ^32,33^. In our study, we show that smoking-induced interstitial myocardial fibrosis is more likely to be present among women when compared to men.

The mechanisms contributing to a greater risk of cardiovascular disease in female smokers compared to their male counterparts are not well understood. However, there appear to be significant sex related differences with regard to susceptibility to heart disease ^34^. In a recent study, Chehab et. al showed that sex hormone imbalances were associated with higher markers of fibrosis by CMR in men. Moreover, higher levels of total testosterone was inversely associated with interstitial myocardial fibrosis in post-menopausal women after controlling for smoking status at baseline ^35^. In postmenopausal women, the loss of estrogen and a shift toward android fat distribution further potentiate smoking’s fibrotic effects, a mechanism less relevant in men, who lack this hormonal axis^36^. In addition, smoking has been shown to differentially increase pro-inflammatory cytokines without enhancement in an inflammatory response in women compared to men^37^ which is known to be associated with increase interstitial myocardial fibrosis^38^.

Previous studies have suggested, that women are more predisposed to concentric cardiac remodeling compared to men, who are more predisposed to eccentric cardiac remodeling ^39^. Although premenopausal women are much less prone to develop cardiovascular disease than men of similar age, this advantage no longer applies after menopause ^40^. The mechanisms contributing to the greater risk of concentric cardiac remodeling in women remain incompletely understood, but there appears to be important sexual dimorphisms in ventricular-vascular structure and function that develop with aging which may predispose to concentric remodeling in women ^41^. In this regard, it has been suggested that increases in the activities of proteolytic enzymes such as matrix metalloproteinases, calpains, cathepsins, and caspases contribute to the process of cardiac remodeling ^42^.

Despite being an established risk factor for cardiovascular disease, cigarette smoking interacts with known risk factors like physical inactivity and obesity, as well as several other cardiovascular pathologic processes to greatly increase the risk for worse cardiovascular outcomes. In our study, cigarette smoking is an important marker of myocardial fibrosis among women which remained in fully adjusted model, when compared to male smokers.

Finally, although the absolute magnitude of the association between cigarette smoking and myocardial fibrosis, as measured by ECV and native T1 time, appears to be modest (e.g., a 0.1% increase per cigarette smoked per day or approximately 2% per pack per day), the critical finding lies in the marked sex-based difference in this relationship. Our results demonstrate that smoking is significantly associated with increased myocardial fibrosis markers in women, but not in men, underscoring a potential sex-specific vulnerability. This divergence, further illustrated in the Figure 1 through the differential trends in ECV and native T1, highlights the1 underlying differences in the pathophysiology of cardiovascular disease between sexes. These findings support the growing body of evidence suggesting that women may have heightened myocardial sensitivity to external stressors such as smoking, which may partly explain their distinct clinical presentation and progression of cardiovascular disease.

## LIMITATIONS

Given the cross-sectional study design, a causal relationship between smoking and myocardial fibrosis cannot be established. In addition, more than 95% of the female participants in this study were post-menopausal thus the association between smoking and myocardial fibrosis in female participants can only be applied to post-menopausal women. In addition, native T1 and ECV might be increased due to other pathologies. We could not control for viral infections, which is known to increase IMF, as the data is only available at Exam 1. Similarly, C-Reactive Protein data was only available at baseline.

## CONCLUSION

In this multiethnic, population-based study, cigarette smoking is associated with higher CMR measures of interstitial myocardial fibrosis (reflected by higher ECV and longer native T1 time), in women but not in men. This indicates differences in the pathophysiology of cardiovascular diseases by sex. However, additional studies are required to further understand such important relationships.

## Data Availability

All data produced in the present study are available upon reasonable request to the authors.

## Acknowledgments

The authors thank the other investigators, the staff, and the participants of the MESA study for their valuable contributions. A full list of participating MESA investigators and institutions can be found at http://www.mesa-nhlbi.org. The views expressed in this article are those of the authors and do not necessarily represent the views of the National Heart, Lung, and Blood Institute; the National Institutes of Health; or the U.S. Department of Health and Human Services.

## ABBREVIATIONS AND ACRONYMS

CMR: Cardiac Magnetic Resonance
ECV: Extracellular Volume
LGE: Late Gadolinium Enhancement
IMF: Interstitial-Myocardial-Fibrosis
nT1: Native T1 Time
CVD: Cardiovascular D1isease

---

**Supplemental Figure 1: Inclusion and Exclusion Criteria**

