## Supplementary figures and images for "Sex-Difference of Associations between Cigarette Smoking and Myocardial Fibrosis: The Multi-Ethnic Study of Atherosclerosis"

### Supp Fig 1

#### Supplemental Figure 1: Inclusion and Exclusion Criteria

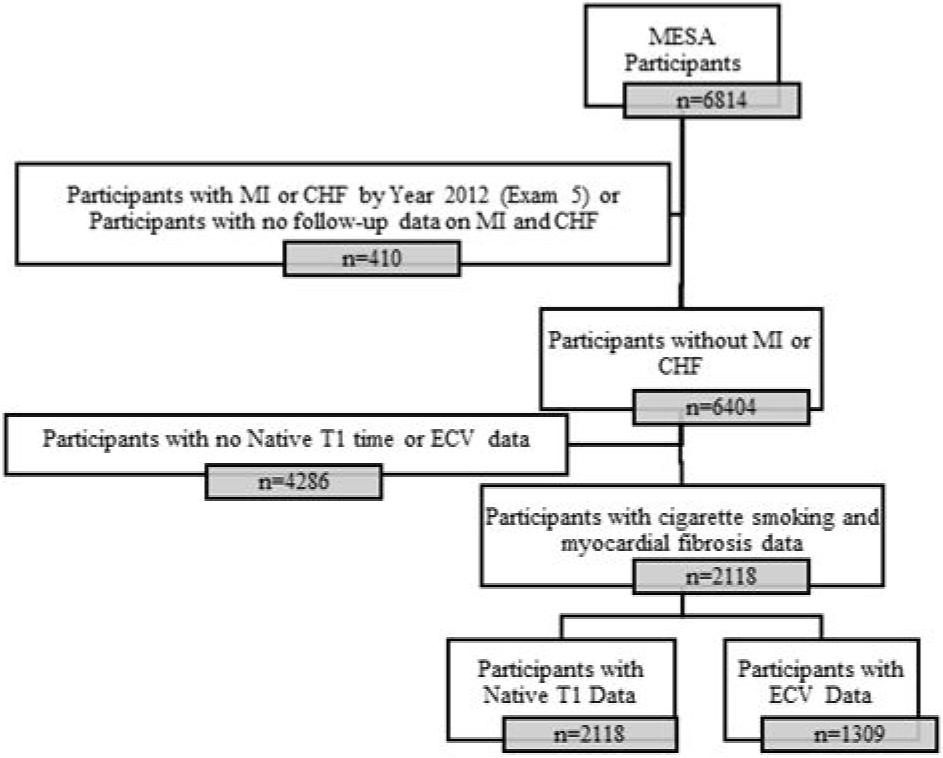
